# Field assessment of BinaxNOW antigen tests as COVID-19 treatment entry point at a community testing site in San Francisco during evolving omicron surges

**DOI:** 10.1101/2022.08.17.22278913

**Authors:** John Schrom, Carina Marquez, Chung-Yu Wang, Aditi Saxena, Anthea M. Mitchell, Salu Ribeiro, Genay Pilarowski, Robert Nakamura, Susana Rojas, Douglas Black, Maria G. Contreras Oseguera, Edgar Castellanos Diaz, Joselin Payan, Susy Rojas, Diane Jones, Valerie Tulier-Laiwa, Aleks Zavaleta, Jacqueline Martinez, Gabriel Chamie, Carol Glaser, Kathy Jacobsen, Maya Petersen, Joseph DeRisi, Diane Havlir

**Author notes:** Corresponding author: Diane Havlir, MD, University of California, San Francisco, 995 Potrero Avenue, CA. These authors contributed equally to this work (Joseph DeRisi, Diane Havlir).

## Abstract

**Background:** COVID-19 oral treatments require initiation within 5 days of symptom onset. Although antigen tests are less sensitive than RT-PCR, rapid results could facilitate entry to treatment. As SARS-CoV-2 variants and host immunity evolve, it is important to characterize the use case for rapid antigen tests for treatment entry.

**Methods:** We collected anterior nasal swabs for BinaxNOW and RT-PCR testing and clinical data at a walk-up, community site in San Francisco, California between January and June 2022. SARS-CoV-2 genomic sequences were generated from positive samples and classified according to subtype and variant. Monte Carlo simulations were conducted to estimate the expected proportion of SARS-CoV-2 infected persons who would have been diagnosed within 5 days of symptom onset using RT-PCR versus BinaxNOW testing.

**Results:** Among 25,309 persons tested with BinaxNOW, 2,952 had concomitant RT-PCR. 1321/2952 (44.7%) were SARS-CoV-2 RT-PCR positive. We identified waves of predominant omicron BA.1, BA.2, BA.2.12, BA.4, and BA.5 among 720 sequenced samples. Among 1,321 RT-PCR positive samples, 938/1321 (71%) were detected by BinaxNOW; 95% (774/817) of those with Ct value <30 were detected by BinaxNOW. BinaxNOW detection was consistent over lineages. In analyses to evaluate entry to treatment, BinaxNOW detected 82.7% (410/496, 95% CI: 79-86%) of persons with COVID-19 within 5 days of symptom onset. In comparison, RT-PCR (24-hour turnaround) detected 83.1% (412/496 95% CI: 79-86%) and RT-PCR (48-hour turnaround) detected 66.3% (329/496 95% CI: 62-70%) of persons with COVID-19 within 5 days of symptom onset.

**Conclusions:** BinaxNOW detected high viral load from anterior nasal swabs consistently across omicron sublineages emerging between January and June of 2022. Simulations support BinaxNOW as an entry point for COVID-19 treatment in a community field setting.

## INTRODUCTION

SARS-CoV-2 rapid antigen tests are public health tools that can be used for COVID-19 diagnosis, to identify persons most at risk for transmission, and as mileposts for isolation [1]. They effectively detect high levels of virus but are less sensitive than reverse transcriptase polymerase chain reaction (RT-PCR) assays at lower viral levels. Rapid antigen tests are used as a surrogate for infectiousness based on their correlation to in vitro cultures, although this application is imperfect with gaps in existing data [2-6]. Rapid antigen tests have the advantage of providing results within minutes to hours and can be done outside of medical clinics or in the home [1, 7]. One of the limitations of these assays is that during the upswing of virus after initial infection, they detect fewer cases compared to RT-PCR testing [8]. For this reason, repeat rapid antigen testing is recommended for persons with suspected infection in the case of a negative rapid antigen test.

There are now 2 oral treatments—nirmatrelvir/ritonavir (Paxlovid™) and molnupiravir (Lagevrio™)—for persons with COVID-19 deemed at high risk for progression to serious disease [9]. Both treatments require initiation within 5 days of COVID-19 symptom onset. Given that delay times for the return of PCR test results vary, but are rarely less than 24 hours, rapid antigen tests could play an important role in identifying persons who could benefit from treatment; however, it remains unclear to what extent this advantage outweighs the inherent limitations of sensitivity in the earliest stages of infection.

We have been operating a walk-up COVID-19 rapid test-and-respond site in the Mission District of San Francisco through an academic, community, and public health partnership that provides low-barrier services that extend the reach of traditional health systems and conducts ongoing molecular surveillance for SARS-CoV-2 [10-12]. The program includes community-led provision of education and supplies for low-income households with new COVID-19 diagnoses [13]. We have previously described the performance of the BinaxNOW rapid antigen tests during various surges of the COVID-19 pandemic in this setting; for example, we demonstrated that rapid tests can be used as a public health tool to increase effective isolation time due to the rapid turnaround of results linked to a community-led response team [11, 14].

Since January 2022, the community served at our testing site has experienced omicron surges with omicron waves of BA.1, BA.2, BA.2.12, BA.4 and now BA.5 sublineages. With the objective of enabling persons in the community with COVID-19 optimal access to new oral treatment within the 5-day window from symptom onset, we sought to evaluate BinaxNOW compared to standard RT-PCR in our community setting for case detection for treatment evaluation. With reports of varying symptom presentations with COVID-19 variants and the evolving immunity of the population due to prior infection, vaccination, or both, we also evaluated BinaxNOW compared to RT-PCR for case detection by time from symptom onset for SARS-CoV-2 sublineages and over the six-month period of January to June 2022.

## METHODS

### Study Setting and Testing Procedures

The Unidos en Salud testing site is a walk-up, high volume, outdoor, free, rapid COVID-19 testing and vaccine site located in a parking lot on the major commercial walkway in the Mission District of San Francisco, California. The site offers weekend and weekday hours and serves a predominant Latino and immigrant community with a high proportion of frontline workers and multi-generational families that have experienced a disproportionate burden of COVID in San Francisco [15-17]. The site is led by an academic (University of California, San Francisco and Chan Zuckerberg Biohub), community (Latino Task Force), and public health (San Francisco Department of Public Health) partnership.

Prior to testing, persons provide demographic characteristics, type and onset of symptoms, vaccine status, and informed consent. Certified laboratory assistants collect bilateral anterior nasal swabs for BinaxNOW according to manufacturer specification. They collected a second swab for RT-PCR in a DNA/RNA Shield (Zymo research) [18]. To assess BinaxNOW performance during times of different circulating sublineages, we submitted samples for RT-PCR on all specimens during two periods: January 1 to 31 (“January” period) and May 14 to June 25 (“June” period). For the remainder of the time, as part of ongoing surveillance, RT-PCR was submitted for sequencing for BinaxNOW positive samples only. Certified readers with oversight by a quality control manager read BinaxNOW cards. We returned results to clients using secure messaging in the Primary Health platform within an hour. All persons with a positive test were contacted within 2 hours by bilingual staff to provide guidance and offer support services.

### SARS-CoV-2 genomic sequencing

As previously described, we performed RT-PCR using N and E gene probes on a random sample of the nasal swabs in the DNA/RNA shield with a positive human control (RNase P) [18]. The assay limit of detection is 100 viral copies per milliliter. Cycle threshold below 40 are considered positive. For samples in January, March, April, May, and June 2022, the Illumina platform (NextSeq/NovaSeq) and the ARTIC Network V3 amplicon strategy were used for full viral genome sequencing as described [19]. Consensus genomes were assembled using the freely available CZID pipeline (czid.org), and variant lineages were called using the Pangolin Lineage Assigner (https://pangolin.cog-uk.io). Complete genomes were deposited in GISAID (virus name “hCoV-19/USA/CA-UCSF-JDxxxx/2022”).

### Analyses

We calculated the proportion of persons testing BinaxNOW positive among all persons testing positive on RT-PCR and stratified analyses by SARS-CoV-2 lineage and time from symptom onset. Monte Carlo simulations were then conducted to estimate the expected proportion of participants that would have been identified as positive within five days of symptom onset, and thus eligible for oral treatment evaluation, based on results of RT-PCR versus BinaxNOW testing under varying assumptions on RT-PCR turnaround time and probability of BinaxNOW repeat testing. The population for this set of analyses included all symptomatic participants positive for SARS-CoV-2 by RT-PCR between January and June 2022 with symptom onset in the past five days. We simulated hypothetical time to return of RT-PCR results based on a exponentially distributed random variable. Participants who tested negative on an initial BinaxNOW test were assigned a probability of testing positive on serial test in three days based on a binomially distributed random variable. For the main analysis, 1000 bootstrapped samples were taken using an expected RT-PCR return time of 1 day and expected return following initial negative BinaxNOW test of 15%. Sensitivity analyses were conducted by varying the expected RT-PCR return time among 1 hour, 1 day, 1.5 days, and 2 days, and varying the expected BinaxNOW return probability from 0%, 15%, 25%, 50%, 75%, and 100%. Participants were considered to have been eligible for evaluation for oral treatment by BinaxNOW if they either: 1) initially tested positive, or 2) initially tested negative, were expected to return for testing, and their return would be within five days of symptom onset. Participants were considered to have been eligible by RT-PCR if their expected return of results fell within five days of symptom onset. Results are reported with 95% confidence intervals based on the bootstrapped samples.

#### Ethics statement

The UCSF Committee on Human Research determined the study met criteria for public health surveillance. All participants provided informed consent for dual testing.

## RESULTS

We evaluated data from 25,309 visits between January 1 and June 26, 2022 with valid results from BinaxNOW. During the entire study period, 6,672 (26%) of visits had a positive BinaxNOW test (Supplementary Figure 1). During the January and June periods, 2,952 of these persons had valid concomitant RT-PCR results and BinaxNOW tests. Among these 2,952 persons, 50.1% were self-reported as female, 47.2% as male, and 2.7% as other (Table 1). In this population, 74.6% self-reported as Latinx or American Indian from Central or South America. There were 1,321/2,952 (45%) SARS-CoV-2 RT-PCR positive samples.

**Table 1.**
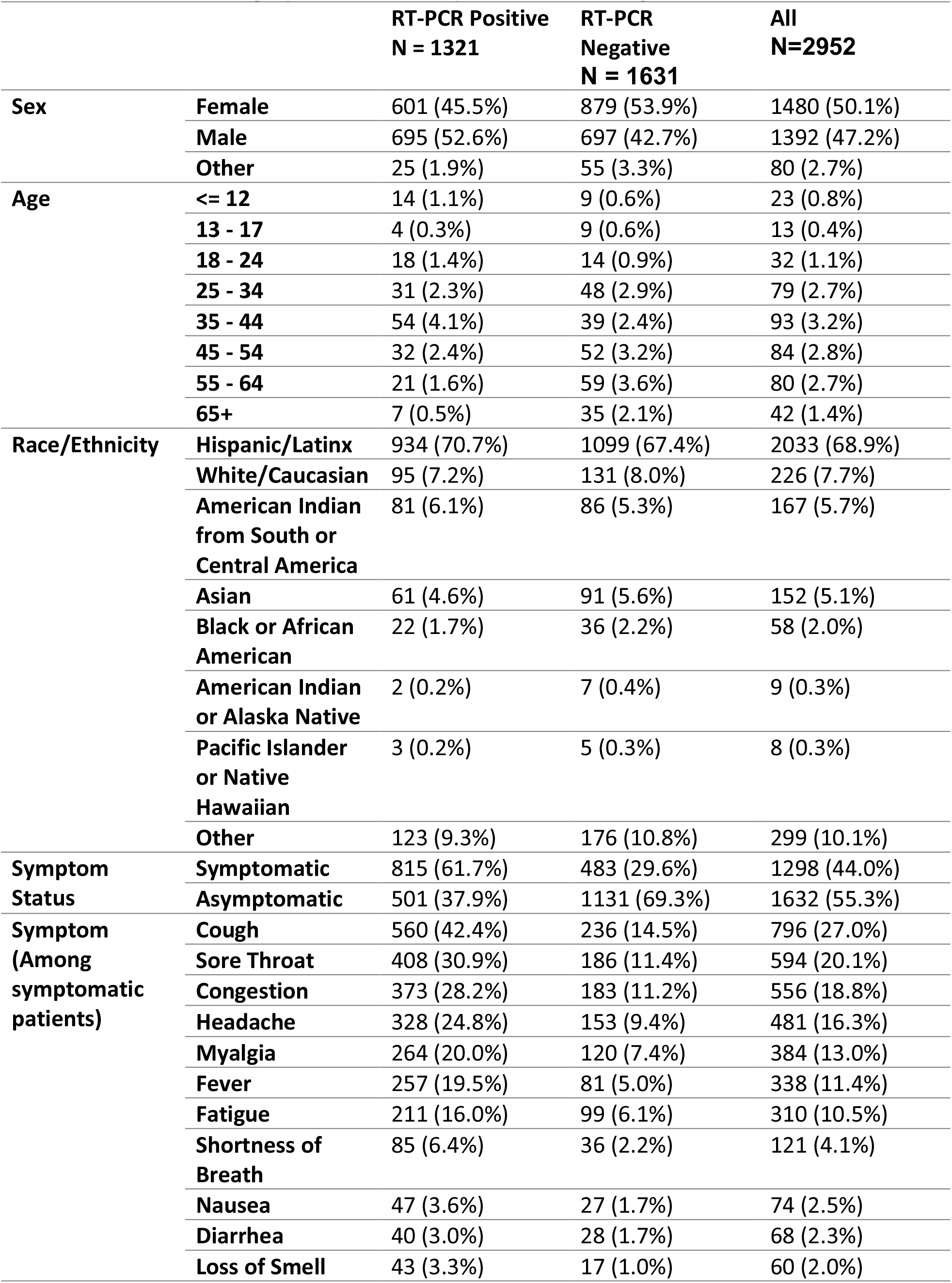
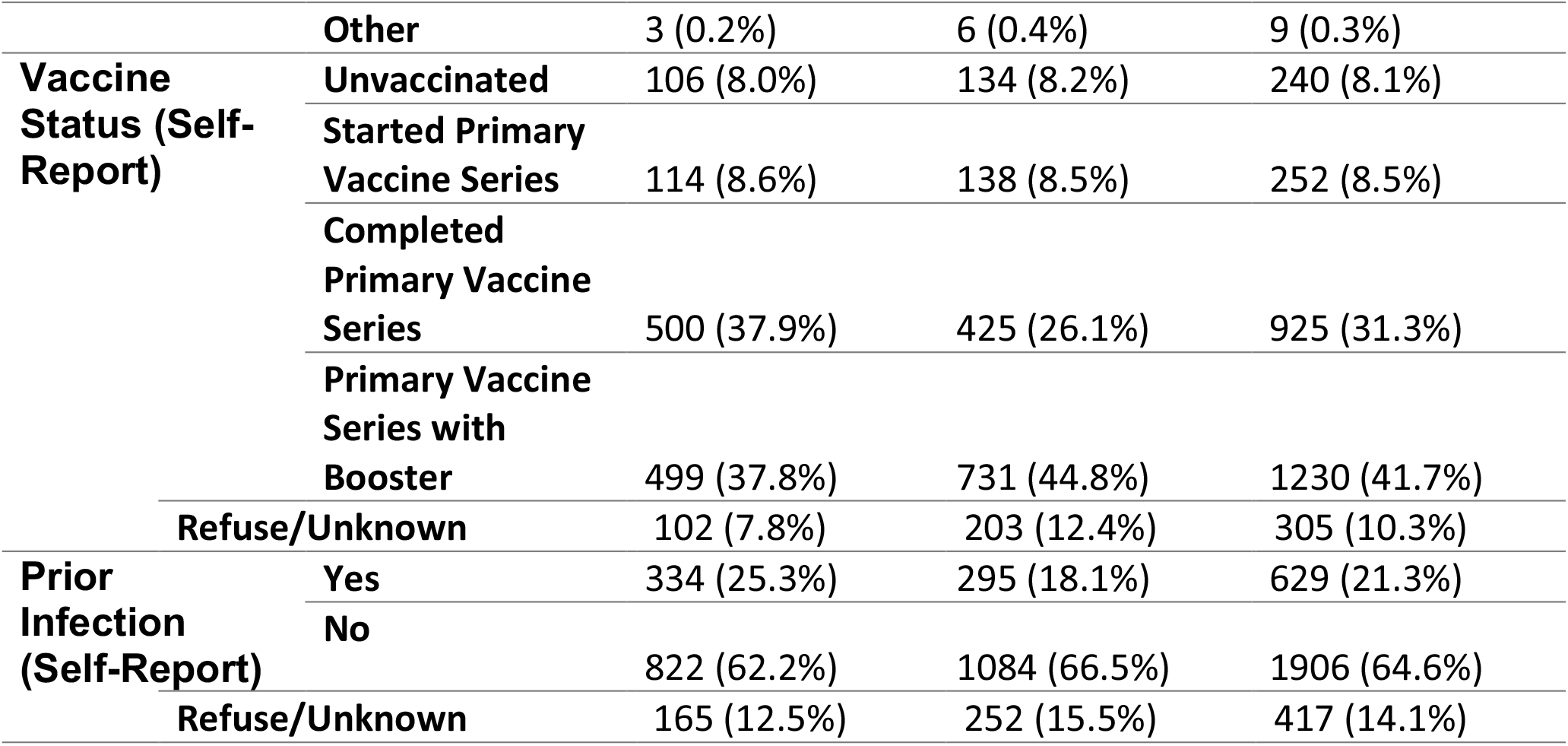
Baseline Demographics for Participants, stratified by Nasal Swab RT-PCR result.

Of the 1,321 persons with positive RT-PCR, 815 (62%) reported at least one symptom associated with COVID-19. The most commonly reported symptoms were cough (42.4%), sore throat (30.9%) and congestion (28.2%). As reported previously, loss of smell was rare (3%) [20]. Primary vaccine series only was reported by 31.3% overall and at least one booster by 41.7%. Among those RT-PCR positive, 37.9% had received the primary series only, 32.9% had one booster and 4.9% had 2 boosters; 25.3% reported prior infection; and 23.8% reported vaccination plus at least one prior infection. Prior vaccine and infection status was by self-report.

We sequenced 720/6,672 (10.8%) of the specimens between January and June 30. Over 98% of the sequences were omicron. In January, the predominant omicron lineage was BA.1; BA.2 became predominant by the end of March. In May, BA.4 and BA.5 appeared, with BA.5 making up 33% of sequenced samples by the end of the study period.

Among 1,321 RT-PCR positive samples, 71% were detected by BinaxNOW; 95% (774/817) of RT-PCR positive sample with Ct value below 30 were detected by BinaxNOW. BinaxNOW detected 65% (443/686) and 77% (490/635) of all RT-PCR positives in January and June, respectively (Figure 2). BinaxNOW detected over 90% of RT-PCR positive samples with Ct < 30 across each sublineage (Supplementary Figure 2). When we examined BinaxNOW positivity stratified by time from symptom onset regardless of Ct, BinaxNOW positivity was 177/216 (82%) in first 2 days compared to 360/448 (80%) for days 3-10. However, in January BinaxNOW detected 80/110 (73%) of RT-PCR positive in days 1-2 from symptom onset compared to 97/106 (92%) in June (Figure 3). BinaxNOW detection of RT-PCR positive cases with Ct < 30 was 90% or more across strata of self-reported vaccine status or prior SARS-CoV-2 infection (Supplementary Figure 3).

**Figure 1.**
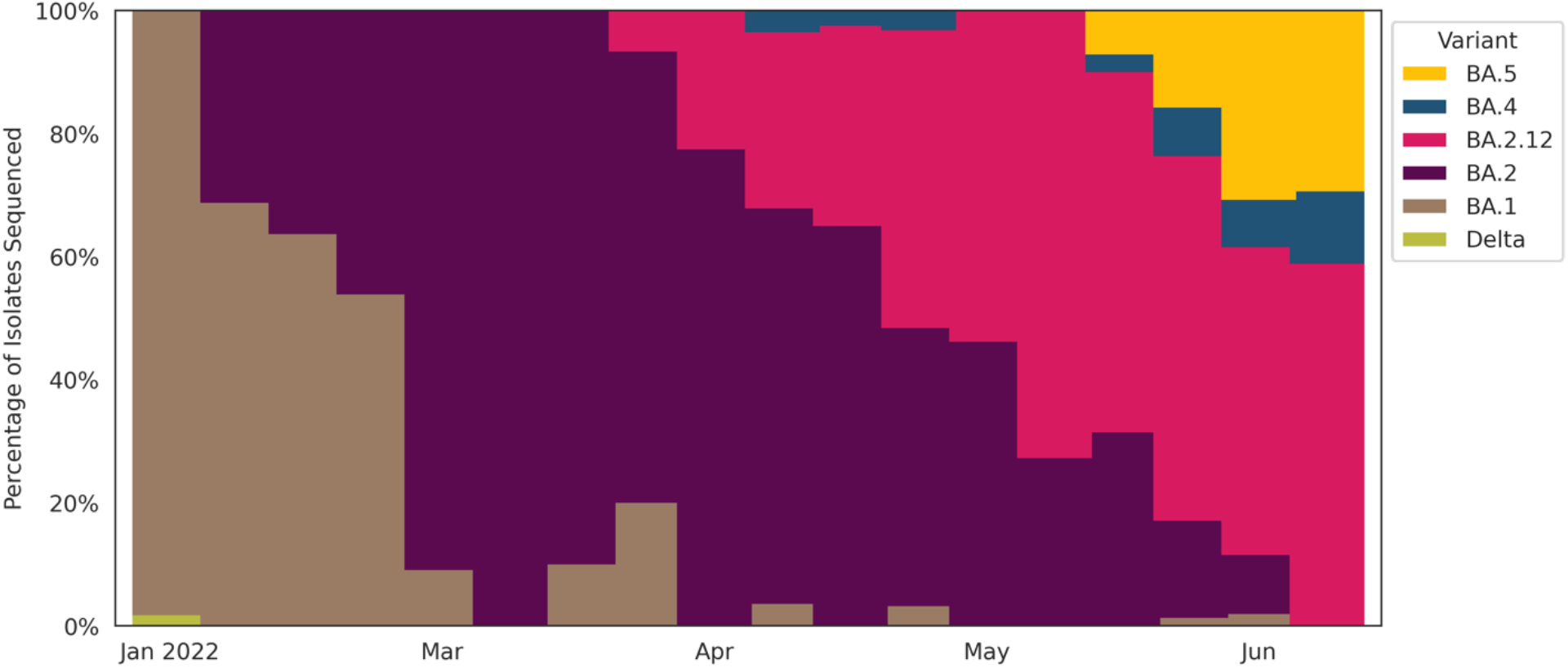
SARS-CoV-2 Lineages of all RT-PCR or rapid antigen test (BinaxNOW) positive participants sequenced between January 1 and June 26, 2022. Each vertical bar corresponds with one week. The height of each color segment within the bar corresponds with the percent of participants in that week whose sequences were identified as that color’s lineage. Abbreviations: Ct, cycle threshold; RT-PCR, reverse transcriptase polymerase chain reaction.

**Figure 2.**
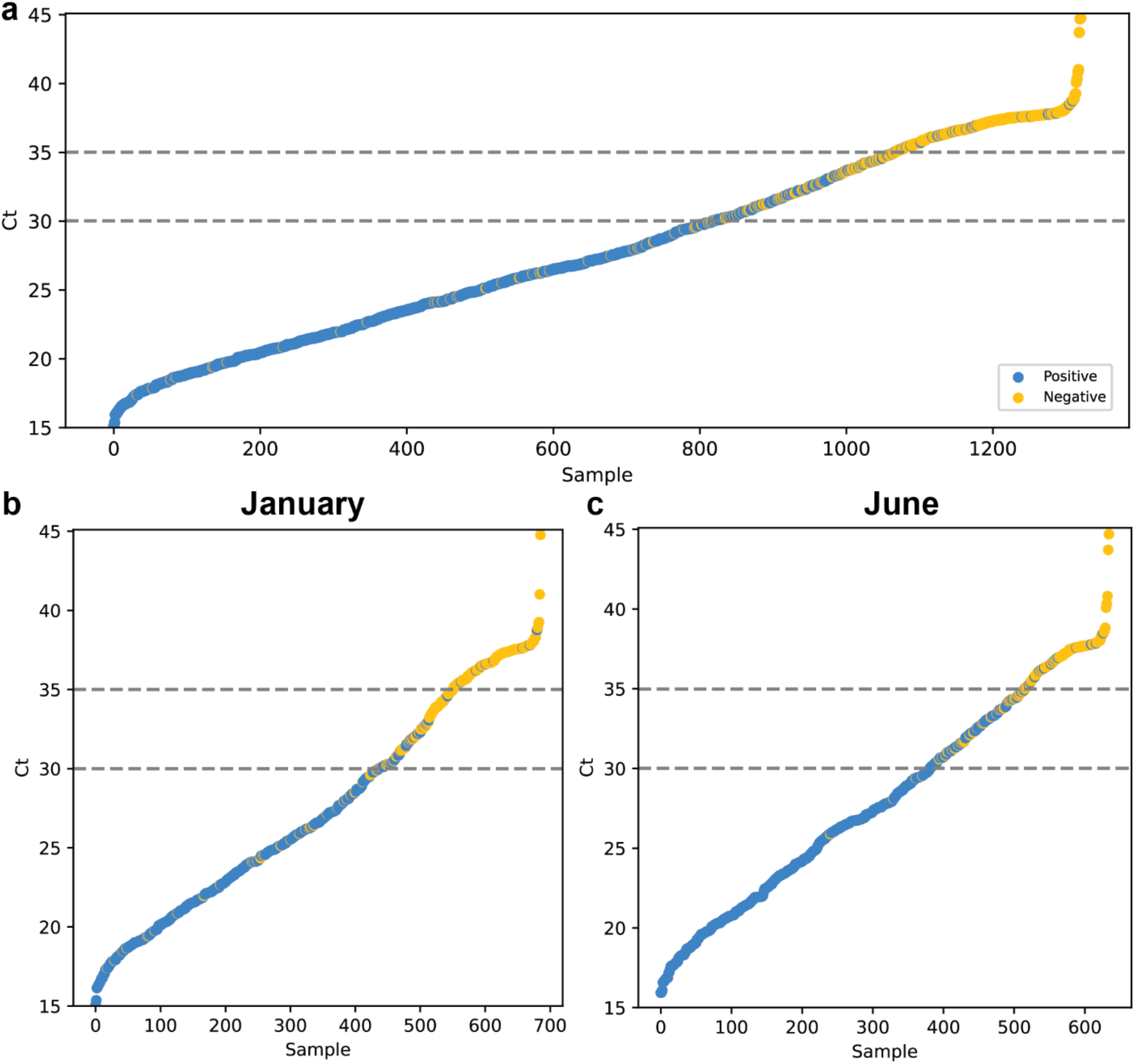
SARS-CoV-2 RT-PCR Ct values and BinaxNOW rapid antigen test results of all RT-PCR positive participants tested between January 1 and June 26, 2022 (panel a) and stratified according to January or June test date (panels b and c). Average viral Ct values of all individuals with positive RT-PCR test results (N=1321) are plotted in ascending order of Ct value. Each point represents one individual. Blue circles are individuals whose samples were positive on both rapid antigen test (BinaxNOW) and on RT-PCR test. Orange circles represent individuals who were RT-PCR positive but rapid antigen test negative. Abbreviations: Ct, cycle threshold; RT-PCR, reverse transcriptase polymerase chain reaction.

**Figure 3.**
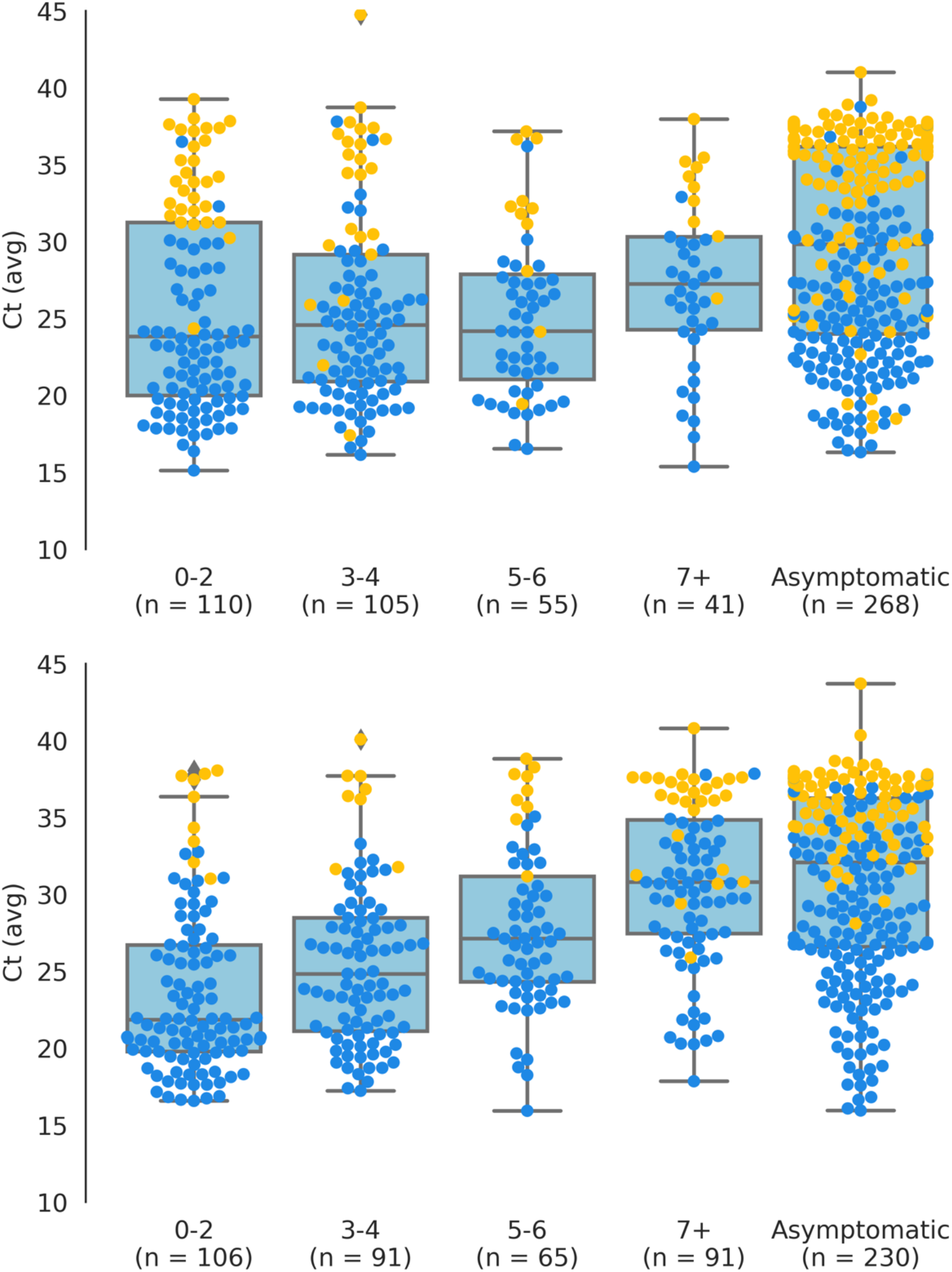
SARS-CoV-2 RT-PCR Ct values and BinaxNOW rapid antigen test results of participants tested between January 1 and June 26, 2022, stratified by days since symptom onset for the January (a) and June (b) periods. Box plot shows first quartile, median, and third quartile in the shaded region; diamonds indicate outliers beyond 1.5 times the interquartile range. Blue circles overlaid are individuals whose samples were positive on both rapid antigen test (BinaxNOW) and on RT-PCR test. Orange circles represent individuals who were RT-PCR positive but rapid antigen test negative. Abbreviations: Ct, cycle threshold; RT-PCR, reverse transcriptase polymerase chain reaction.

In analyses to assess the potential of each testing modality to identify persons eligible for treatment evaluation, BinaxNOW detected 82.7% (410/496, 95% CI: 79-86) of RT-PCR positive persons presenting within the 5-day window from symptom onset. In comparison, RT-PCR (assuming a 24-hour turnaround) would have detected 83.1% of eligible persons within the 5-day window (412/496 95% CI: 79%,86%) and RT-PCR (assuming a 48-hour turnaround) would have detected 66.3% (329/496 95% CI: 62-70) (Table 2). In Monte Carlo simulations to estimate the expected percent of participants that would have been identified as eligible for oral treatment evaluation based on results of SARS-CoV-2 RT-PCR versus BinaxNOW, in January, there was no difference between the two testing modalities (Figure 4). In June, BinaxNOW would have identified 86.1% (95% CI 81-91%) while RT-PCR (assuming 24-hour turnaround) would have identified 71.5% (95% CI 55-84%), an absolute difference of 14.7% (95% CI 8.7-20.7) favoring BinaxNOW. Simulations evaluating differences between the 2 assays using a range of RT-PCR turnaround times, and probability that persons with negative BinaxNOW would return for a repeat test (Supplementary Figure 4) found that, in settings where RT-PCR results can be generated and provided to clients within hours, RT-PCR would identify a higher proportion of persons eligible for treatment evaluation. However, under a range of assumptions that reflect common settings in community sites, such as where our study was conducted, BinaxNOW identified as many or more persons eligible for treatment evaluation than would have been evaluated with RT-PCR due to the reduced turnaround time of rapid antigen tests.

**Table 2.** Results of analysis comparing BinaxNOW to RT-PCR test under multiple assumptions.

| **Testing Method** | **Assumption** | **Positive within 5 days (%)** | **95% CI** | **Mean Time to Positive (days)** |
| --- | --- | --- | --- | --- |
| **BinaxNOW** | **Return for repeat testing** | 90.5% | 88 – 93% | 3.1 |
|  | **No return for repeat testing** | 82.7% | 79 – 86% | 2.9 |
| **PCR** | **24hr turnaround** | 83.1% | 79 – 86% | 3.5 |
|  | **48hr turnaround** | 66.3% | 62 – 70% | 4.1 |
|  | **72hr turnaround** | 43.5% | 39 – 48% | 4.7 |

**Figure 4.**
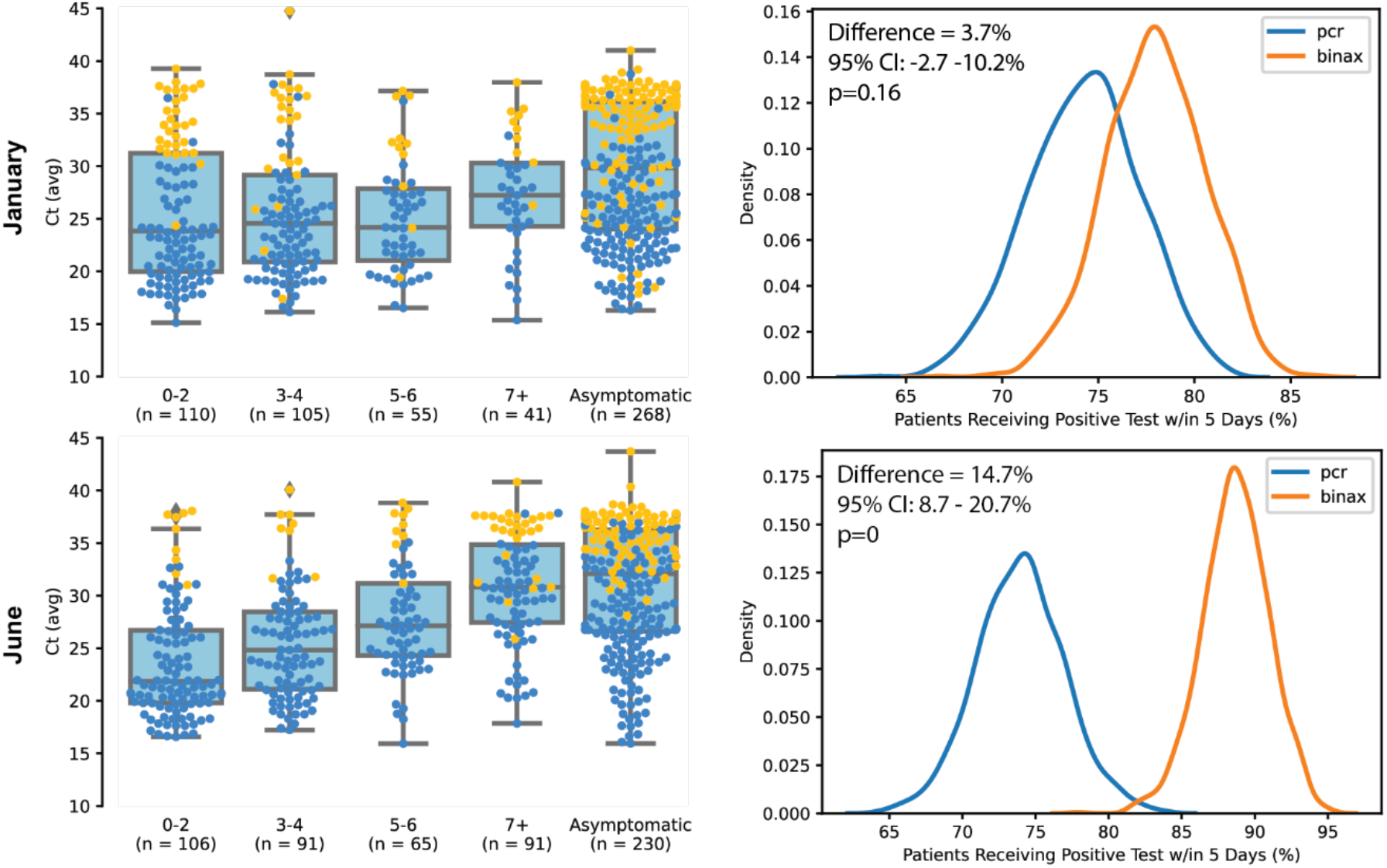
Results of Monte Carlo simulations to estimate the expected percent of participants that would be eligible for oral treatment evaluation based on results of SARS-CoV-2 RT-PCR or rapid antigen test (BinaxNOW) testing under different scenarios. Each row corresponds to a different scenario; the top row uses conditions from January 2022 and the bottom row uses conditions from June 2022. The left column shows the distribution of SARS-CoV-2 RT-PCR Ct by days since symptom onset used as input into the simulations; orange dots correspond to participants who were RT-PCR positive but rapid antigen test negative and blue dots correspond to participants who were both RT-PCR and rapid antigen test positive. The right column show the resulting distributions of percent of participants who would be eligible for oral treatment evaluation based on RT-PCR (blue) versus rapid antigen test (orange) testing. These simulations assume 15% of participants with false negative rapid antigen tests will return and test positive three days after their initial test, and that RT-PCR test results will return after an average of 1 day. The mean difference, 95% confidence interval, and p-value are overlaid on the respective graphs, all calculated using the Monte Carlo simulations. Abbreviations: Ct, cycle threshold; RT-PCR, reverse transcriptase polymerase chain reaction.

## DISCUSSION

We found that the BinaxNOW rapid antigen test detected 71% of RT-PCR positive SARS-CoV-2 anterior nasal swab specimens and 95% of specimens with high levels of RT-PCR positive SARS-CoV-2 (Ct < 30) among persons presenting in this setting where there were high rates of ongoing community transmission and evolving omicron variants. BinaxNOW—even with its lower sensitivity compared to RT-PCR—detected infection among symptomatic persons within the window of five days from symptom onset for treatment eligibility in as many or more cases as RT-PCR assuming a 24- or 48-hour turnaround for PCR results. These data support BinaxNOW as a compliment to RT-PCR testing entry points for identification of individuals in need of treatment evaluation.

Among the many challenges of the public health response to COVID-19 is the need to ensure that tools and guidelines are suitable for rapidly shifting virus and host immunity [21]. To date, SARS-CoV-2 antigen assays, despite having lower sensitivity for viral detection than RT-PCR, have proven an important resource for rapid detection of persons with high levels of virus [1]. However, there is heterogeneity in assays, and there have been mixed reports on changes of rapid antigen performance for detection of omicron variants from in vitro and clinical studies [21-27]. There was even formal recommendations against use of certain antigen tests for omicron detection by the US FDA in 2022 [28]. Together, these reports emphasize the important role of in vitro and field testing for detection of SARS-CoV-2, a virus that continues to circulate and develop new mutations.

Our data can inform the public health utility of only a single assay—BinaxNOW—as assessed in in a community-based setting during recent omicron surges. Although there are many mutations in the SARS-CoV-2 spike mutation in omicron variants, we showed consistently high detection (>90%) for persons with high levels of virus, across omicron sublineages BA.1, BA.2, BA.2.12, and BA.5, using BinaxNOW in RT-PCR positive samples. The detection levels reported here are similar to those we reported for BinaxNOW testing in this community setting when prior SARS-CoV-2 variants were dominating, providing reassurance for use of these tests in the current era [10, 12, 29, 30]. Omicron signature mutations are in the spike protein, but they also in the nucleocapsid protein (NP)—the target of BinaxNOW and many rapid antigen tests. This genetic variability did not appear to affect field performance of BinaxNOW in this study utilizing anterior nasal swab specimens and RT-PCR as a referent.

One of the interesting observations in our data was the lower proportion of RT-PCR samples that were BinaxNOW positive within the first 2 days of symptom onset in January compared to June, a change that appeared to be largely driven by a decrease in viral load at time of testing among persons with recent symptom onset. It has been pointed out previously that rapid antigen assays detect fewer cases than RT-PCR during the upswing of the virus (when viral levels are not yet high enough for rapid antigen detection), and that this is more likely to be seen among testers at the beginning of a surge such as occurred in January [8]. However, this seems unlikely to alone account for the apparent change in the relationship between viral load and symptom onset we observed. It is possible that the viral load at which symptom onset occurs may have increasingly shifted to a lower threshold due to increasing immunity concurrent with the emergence of Omicron. Intriguingly, we observed the opposite temporal pattern here, with an evolution towards higher viral loads immediately following symptom onset between the initial omicron surge in January and six months later, in June of 2022.

There are many possible reasons for these observations. Testing behavior may have changed such that persons reported symptoms and sought testing earlier in January than June. Reinfection, previously an unusual occurrence in the COVID-19 pandemic, was being documented around the world, including among vaccinated persons who may have had lower index of suspicion of COVID-19 [31, 32]. These individuals may have perceived symptoms differently and delayed testing in June compared to January. It is also possible that the variants present in June elicited a pace of inflammatory responses and clinical symptoms in susceptible hosts different than in January. In an elegant modeling exercise using a large data set from England, during the initial days from symptoms onset, variant status had the largest influence on variations in Ct threshold [33]. Finally, it is possible if not probable that a combination of factors contributed.

One of the pillars of the COVID-19 response is oral antimicrobial treatments for those individuals who are at high risk for progression to serious disease [9]. These treatments must be started within 5 days of symptom onset. The fastest entry into identifying persons with suspected COVID-19 for treatment would be a highly sensitive RT-PCR or nucleic acid -based assay with turnaround within hours. However, RT-PCR with rapid turnaround is not accessible in many settings, and in addition, requires a health professional to connect with a patient to convey results and evaluate for treatment. We were interested in evaluating whether in a community field setting with an existing infrastructure to connect with patients, a rapid antigen test such as BinaxNOW, even with its lower sensitivity compared to RT-PCR, could be an important public health tool for treatment evaluation. Our data and simulations with varying distributions of BinaxNOW detection by time from symptom onset show that, over a range of RT-PCR turnaround times plausible in many communities, BinaxNOW identified as many or more persons within a window during which they remained eligible for oral COVID-19 treatment as would have been identified in this window by RT-PCR. Our simulations are not intended to imply that BinaxNOW is a better test than RT-PCR for SARS-CoV-2 detection, but rather to show, using real world data in a community setting, that under a variety of scenarios it is a viable entry point into treatment evaluation.

Our community testing and vaccination site extends the reach of traditional health systems in a public health response to COVID-19 by offering walk-up services that address some of the barriers that persons who are uninsured, monolingual, or managing multiple jobs with nontraditional hours may experience. Although we have shown that BinaxNOW is a reasonable entry point for treatment evaluation in this setting, its impact will require immediate treatment access. At the beginning of the omicron surge, oral treatments for COVID had limited availability. Now that access to oral treatments have increased, we have started a rapid screening and dispensing program for nirmatrelvir/ritonavir for eligible symptomatic individuals testing positive using BinaxNOW. If this approach works, it provides another point-of entry for a community that continues to be disproportionately affected by COVID.

We recognize the limitations of these surveillance data. First, results from our study may not apply to all rapid antigen tests, and home testing using BinaxNOW or other tests are subject to variability in user performance. Second, the comparative performance of these two testing modalities for identifying SARS-CoV-2 infected persons within the 5 day window needed for treatment evaluation will depend on viral burden and time since symptom onset at time of presentation for testing, which in turn may vary across populations, geographical regions, and over time due to factors such as changes in testing behavior, immune landscape, and epidemic context, underscoring the importance of both in vitro and field studies for evolving conditions in various settings. Further, these serial cross-sectional data drawn from a sample of persons presenting for testing do not allow us to distinguish between biological and behavioral explanations for differences in viral load at time of testing for persons with recent symptoms. Third, our measures of BinaxNOW positivity in persons with SARS-CoV-2 on RT-PCR are confined to disease detection by anterior nasal swabs. As has been pointed out by studies obtaining repeated specimens from multiple sites including pharynx, sampling multiple sites can increase disease detection [27]. However, this type of sample ascertainment at high volume community sites would not be feasible. Community low barrier sites such as Unidos en Salud are one part of the public health response and can play an important role in reducing disparities in outbreaks or pandemics fueled by inequities in access to health services.

In conclusion, at a community walk-up testing site, BinaxNOW detected persons with high levels of SARS-CoV-2 infection in multiple omicron sublineages within a time frame enabling an additional entry point for treatment evaluation. Ongoing assessments of assays to facilitate entry points into COVID-19 treatment as the virus, immune responses, and treatment options progress.

## Data Availability

Data are not available

## Acknowledgements

The authors thank the community for their extraordinary support and ongoing partnership.

## Disclaimer

The findings and conclusions in this article are those of the authors and do not necessarily represent the views opinions of the California Department of Public Health or the California Health and Human Services Agency.

---

**Supplemental Figure 1.**
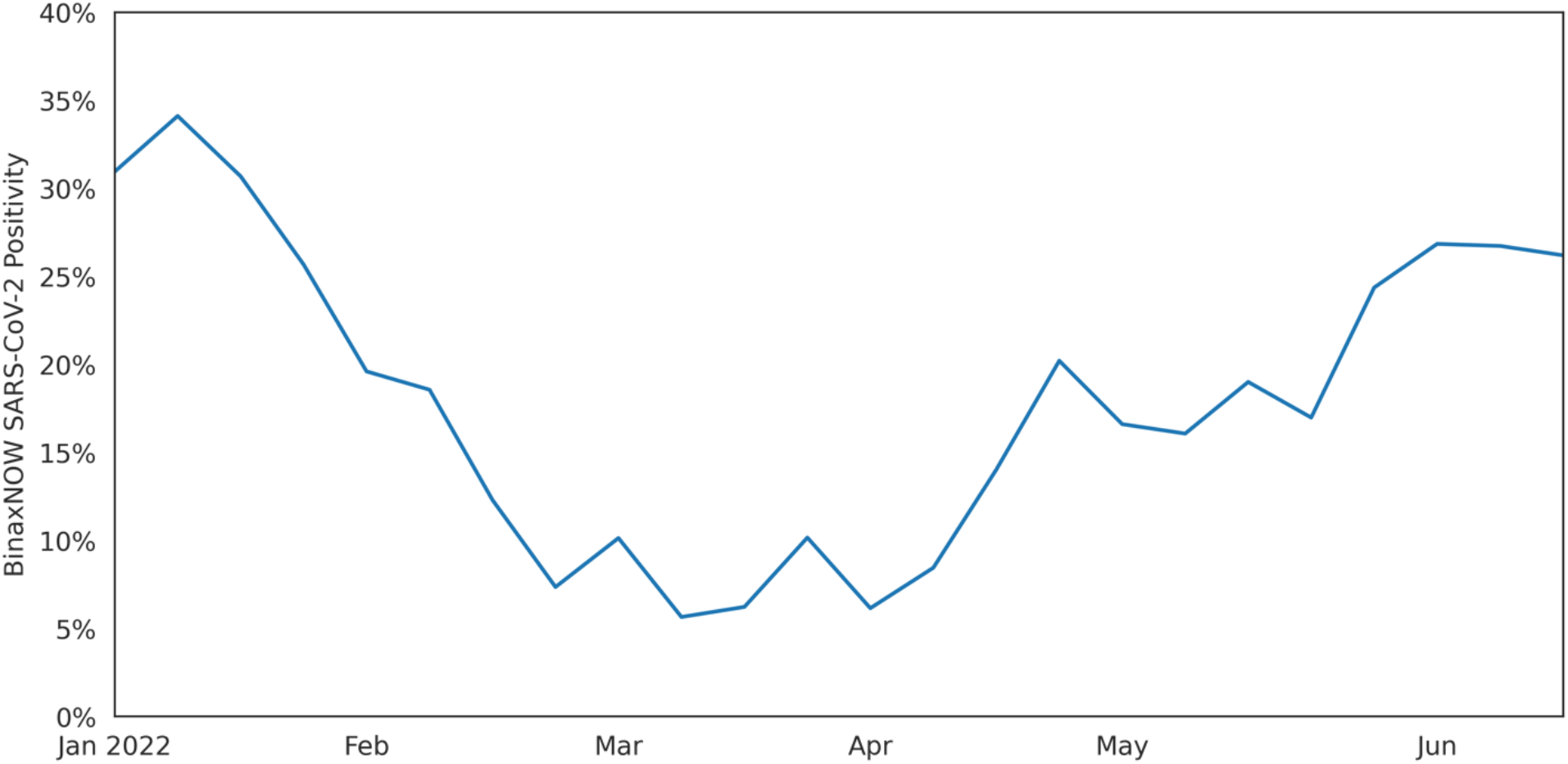
SARS-CoV-2 BinaxNOW rapid antigen test positivity between January 1 and June 26, 2022 at walk-up community testing site Unidos En Salud in the Mission District in San Francisco, California.

**Supplemental Figure 2.**
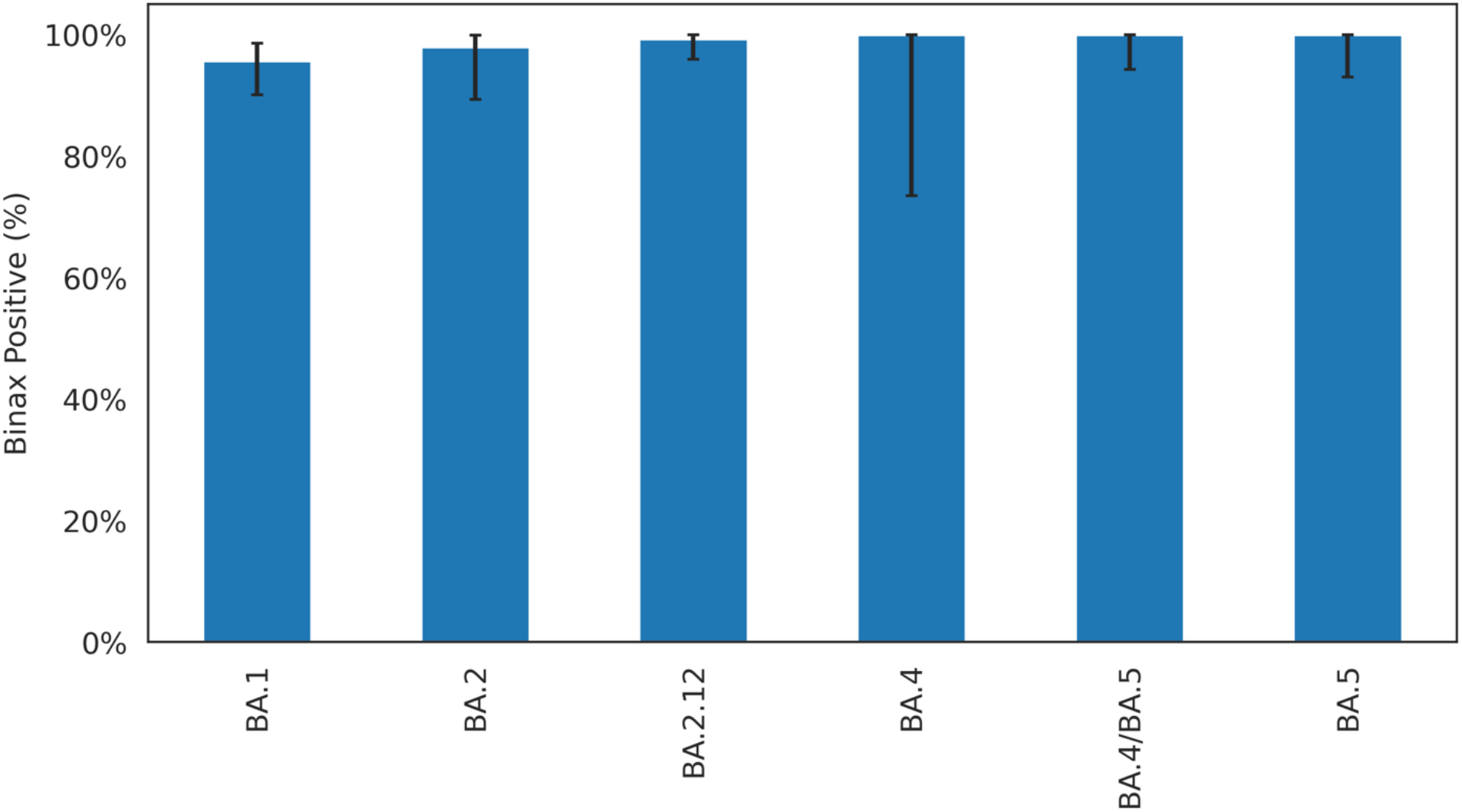
BinaxNOW positivity among SARS-CoV-2 RT-PCR positive participants with Ct < 30, with confirmed lineage data. Each bar represents the BinaxNOW positivity rate among the corresponding lineage and related sublineages, with 95% confidence intervals.

**Supplemental Figure 3.**
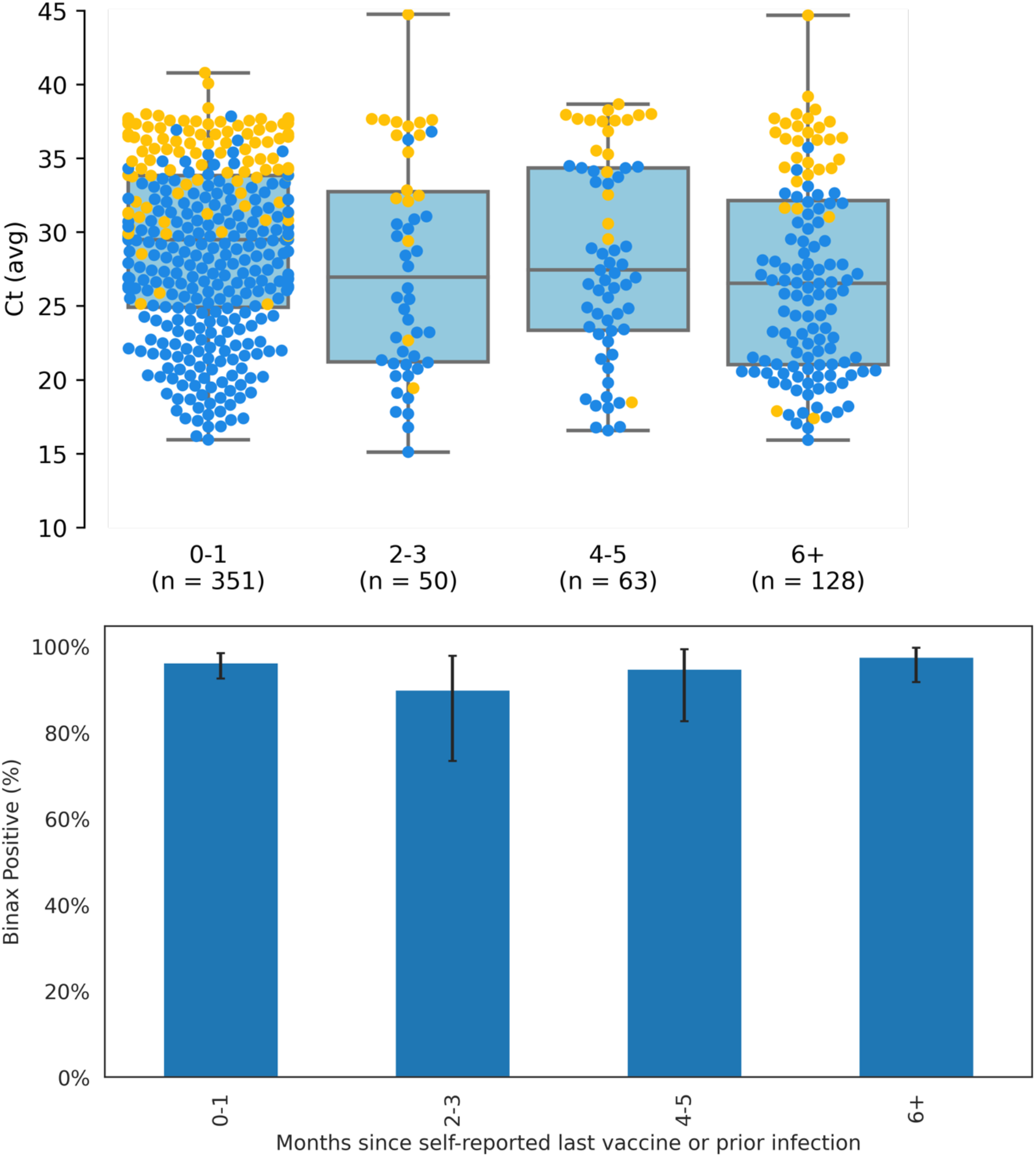
SARS-CoV-2 RT-PCR Ct values and BinaxNOW rapid antigen test results (panel a), and BinaxNOW rapid antigen test detection (panel b) for participants tested between January 1 and June 26, 2022, stratified by months since prior self-reported vaccine or infection status. Panel A shows box plot shows first quartile, median, and third quartile in the shaded region; diamonds indicate outliers beyond 1.5 times the interquartile range. Blue circles overlaid are individuals whose samples were positive on both rapid antigen test (BinaxNOW) and on RT-PCR test. Orange circles represent individuals who were RT-PCR positive but rapid antigen test negative. Panel B shows bar plots with percent of RT-PCR positive participants with SARS-CoV-2 RT-PCR Ct less than 30 who were also BinaxNOW positive. Error bars indicate 95% confidence interval for each estimate. Abbreviations: Ct, cycle threshold; RT-PCR, reverse transcriptase polymerase chain reaction.

**Supplemental Figure 4.**
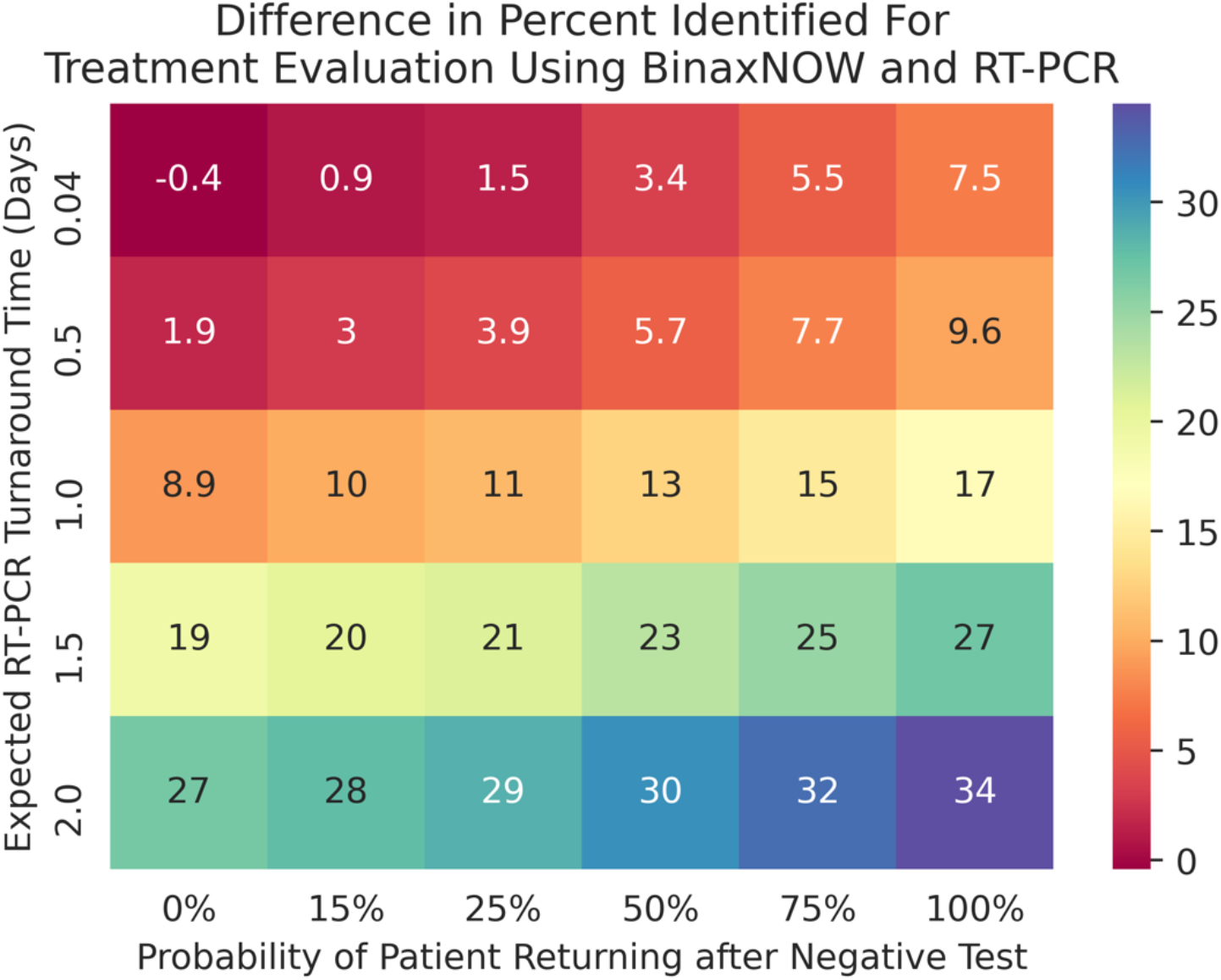
Results of analysis evaluating Monte Carlo simulation sensitivity to underlying assumptions. Rows correspond to variations of the expected time to receive RT-PCR results in days, as modeled by an exponentially distributed random variable; the rows correspond with one hour, half day, full day, day and a half, or two days. The columns correspond with the expected percent of patients returning and testing positive after a false negative, as modeled by a binomially distributed random variable; the columns correspond with 0%, 15%, 25%, 50%, 75%, and 100%. Each cell corresponds with the expected difference between the percent of participants being eligible for oral treatment based on rapid antigen test minus those eligible based on RT-PCR; a positive number indicates rapid antigen test identifying more participants, and a negative number indicated RT-PCR identifying more. Abbreviations: Ct, cycle threshold; RT-PCR, reverse transcriptase polymerase chain reaction.

